# Associations of Midlife Diet Quality with Incident Dementia and Brain Structure: Findings from the UK Biobank Study

**DOI:** 10.1101/2022.05.06.22274696

**Authors:** Jingyun Zhang, Xingqi Cao, Xin Li, Xueqin Li, Xiaoyi Sun, Gan Yang, Meng Hao, Ce Sun, Yang Xia, Huiqian Huang, Terese Sara Høj Jørgensen, George O. Agogo, Liang Wang, Xuehong Zhang, Xiang Gao, Heather Allore, Zuyun Liu

## Abstract

**Objective:** To investigate the associations of midlife diet quality with incident dementia and brain structure.

**Design:** Population-based prospective study and cross-sectional study.

**Setting:** UK Biobank.

**Participants:** In total, 187,783 participants (mean age 56.8 years, 54.9% women) who completed the 24-hour recall dietary questionnaire were included in the prospective study. A subgroup of 25,380 participants (mean age 55.7 years, 52.9% women) with brain structure data were included in the cross-sectional study.

**Main exposure and outcome measures:** Cox proportional hazards models and linear regression models were used to examine the associations of seven diet quality scores, i.e., hPDI (Healthful Plant-based Diet index), MDS (Mediterranean Diet score), aMED (alternate Mediterranean diet), RFS (Recommended Food Score), DASH (Dietary Approaches to Stop Hypertension), MIND (Mediterranean-DASH Intervention for Neurodegenerative Delay diet) and AHEI-2010 (the Alternative Healthy Eating Index-2010), with incident dementia and brain structure (estimated using magnetic resonance imaging), respectively.

**Results:** During a total follow-up of 1,969,993 person-years, 1,363 (0.73%) participants developed dementia. Higher diet quality scores (except for hPDI) were consistently associated with a lower incidence risk of dementia (all P for trend<0.001). For instance, for RFS, the hazard ratios of the intermediate tertile group and the highest tertile group relative to the lowest tertile group were 0.85 (95% confidence interval [95%CI]=0.75 to 0.97) and 0.70 (95%CI=0.61 to 0.81), respectively. Moreover, higher diet quality scores were significantly associated with larger regional brain volumes including volumes of grey matter (GM) in the parietal and temporal cortex and volumes of the hippocampus and thalamus. For instance, higher RFS was associated with larger volumes of GM in the postcentral gyrus (β=16.05±4.08, P<0.001) and the hippocampus (β=5.87±1.26, P<0.001). A series of sensitivity analyses confirmed the main results.

**Conclusion:** Greater adherence to MDS, aMED, RFS, DASH, MIND, and AHEI-2010 were individually associated with lower risk of incident dementia and larger brain volumes in specific regions. This study shows a comprehensive picture of the consistent associations of midlife diet quality with dementia risk and brain health, providing mechanistic insights into the role of healthy diet in the prevention of dementia.

**Summary box:** 

**What is already known on this topic:**

1. Previous prospective studies and meta-analyses suggested significant associations between a few diet quality scores (i.e., MDS, DASH, MIND, and AHEI-2010) and the risk of dementia in different populations; however, the results did not reach agreement.
2. Nutrient intakes or very few diet quality scores have been demonstrated to be associated with brain volumes derived from MRI. There is limited research on the associations of various diet quality scores with the risk of dementia and brain structures in the same population.

**What this study adds:**

1. Greater adherence to MDS, aMED, RFS, DASH, MIND, and AHEI-2010, but not hPDI was individually associated with lower risks of incident dementia.
2. Greater adherence to MDS, aMED, DASH, and AHEI-2010, especially RFS, was individually associated with larger brain volumes in special regions (e.g., parietal and temporal cortex, and hippocampus).
3. This study shows a comprehensive picture of the consistent associations of midlife diet quality with dementia risk and brain health, providing mechanistic insights into the role of healthy diets in the prevention of dementia.

## 1. Introduction

Dementia, mainly including Alzheimer’s Disease (AD) and vascular dementia, is a pivotal public health issue with immense economic burdens at both individual and national levels. According to the World Health Organization, more than 55 million people live with dementia worldwide in 2021, and this number may increase by 10 million per year^1^. Dementia increases the risk of poor patient-centered outcomes, such as cardiovascular diseases^2^, disturbed emotions^3^, and death^4-6^, increases risk of depression among caregivers^7 8^, and limits social interactions^9 10^. Thus, proper preventive and management strategies are critical due to the lack of effective treatments of dementia.

Healthy diet, as a modifiable lifestyle factor, may help prevent incident dementia and dementia progression. Previous prospective studies and meta-analyses suggested significant associations of the following diet quality scores with the risk of dementia: the AHEI-2010 (the Alternative Healthy Eating Index-2010)^11^, DASH (Dietary Approaches to Stop Hypertension)^12^, aMED (alternate Mediterranean diet)^11^, MDS (Mediterranean Diet Score)^13-15^, and MIND (Mediterranean-DASH Intervention for Neurodegenerative Delay Diet)^12^. Yet, nonsignificant associations of MDS^16 17^ and DASH^17^ with dementia have also been reported in different populations. Recently, a healthful Plant-based Diet Index (hPDI) was proposed by Dr. Satija et al^18^; however, no studies have explored the association of hPDI with incident dementia. Currently, there are limited research on the associations of these increasing number of diet quality scores with the risk of dementia in the same population.

Pathological changes in brain structures due to onset or progression of neurodegenerative disorders can be detected using advanced diagnostic techniques including Magnetic resonance imaging (MRI)^19^. Previous studies have demonstrated the potential effects of diet quality or nutrient intakes on brain volumes derived from MRI. The Prospective Investigation of the Vasculature in Uppsala Senior’s (PIVUS) study showed that lower meat intake was associated with greater total brain volume^20^. Results from the community-based Washington Heights/Hamilton Heights Inwood Columbia Aging Project (WHICAP) showed that higher adherence to the Mediterranean diet was associated with less brain atrophy^21^. However, the association of midlife diet quality as assessed by other diet quality scores, such as hPDI, with total brain volumes, and regional volumes of brain, remain largely unknown. The aim of this study was twofold. First, we investigated the prospective associations of midlife diet quality, as measured by seven existing diet quality scores, namely, hPDI, AHEI-2010, DASH, aMED, MDS, MIND, and RFS (Recommended Food Score) with incident dementia using data from the large, population-based UK Biobank Study with a total follow-up time of 1,969,993 person-years (n=187,783). Second, we examined the associations of the above seven diet quality scores with total brain volumes and brain regional volumes from a subgroup of 25,380 participants with complete, high quality MRI images (**Figure 1**).

**Figure 1.**
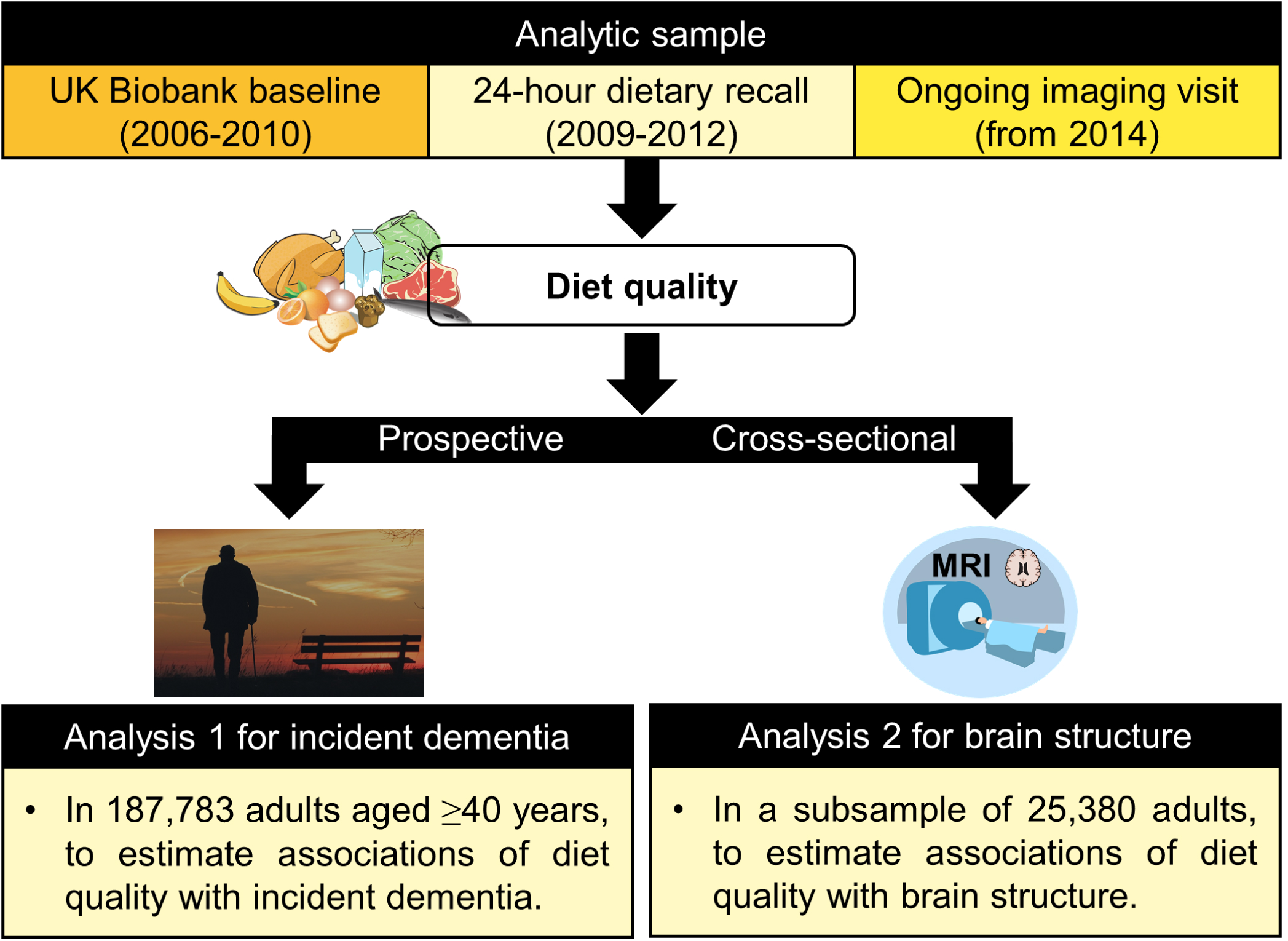
Analytic plan of the study.

## 2. Materials and methods

### 2.1 Study population

The UK Biobank is a large-scale and population-based study that recruited more than half a million participants between 40 and 69 years from 2006 to 2010 in the UK^22^. The UK Biobank collected multi-dimension data, including biological samples (e.g., blood, urine, and saliva), physical measurements data (e.g., blood pressure, weight, and height), questionnaires on health, and genetic data. In addition, it also has invited some original participants back to collect the body, brain, and heart imaging from 2014. More details of UK Biobank are available online (http://www.ukbiobank.ac.uk/). This study included two analytic samples (analysis 1 for incident dementia and analysis 2 for brain structure, see **Figure 1**). First, as shown in **Figure 2**, among 210,971 participants who completed the 24-hour recall dietary questionnaire at least once, 23,188 participants were excluded due to: 1) dementia at baseline (n=75); 2) non-British white (n=9,682, to reduce the ethnic influence on genetic data); 3) lack of APOE genotypes (n=4,169); 4) implausible energy intake (n=4,956, men <800 or >4200 kcal/day, and women <500 or >3500 kcal/day^23^); and 5) the lack of other covariates (n=4306, e.g., education level, smoking status, alcohol consumption), resulting in 187,783 participants (analytic sample 1) included in the prospective study of associations between diet quality and incident dementia. Second, a subgroup of 25,380 participants (analytic sample 2) with brain MRI data was included in the cross-sectional study of associations between diet quality and brain volumes.

**Figure 2.**
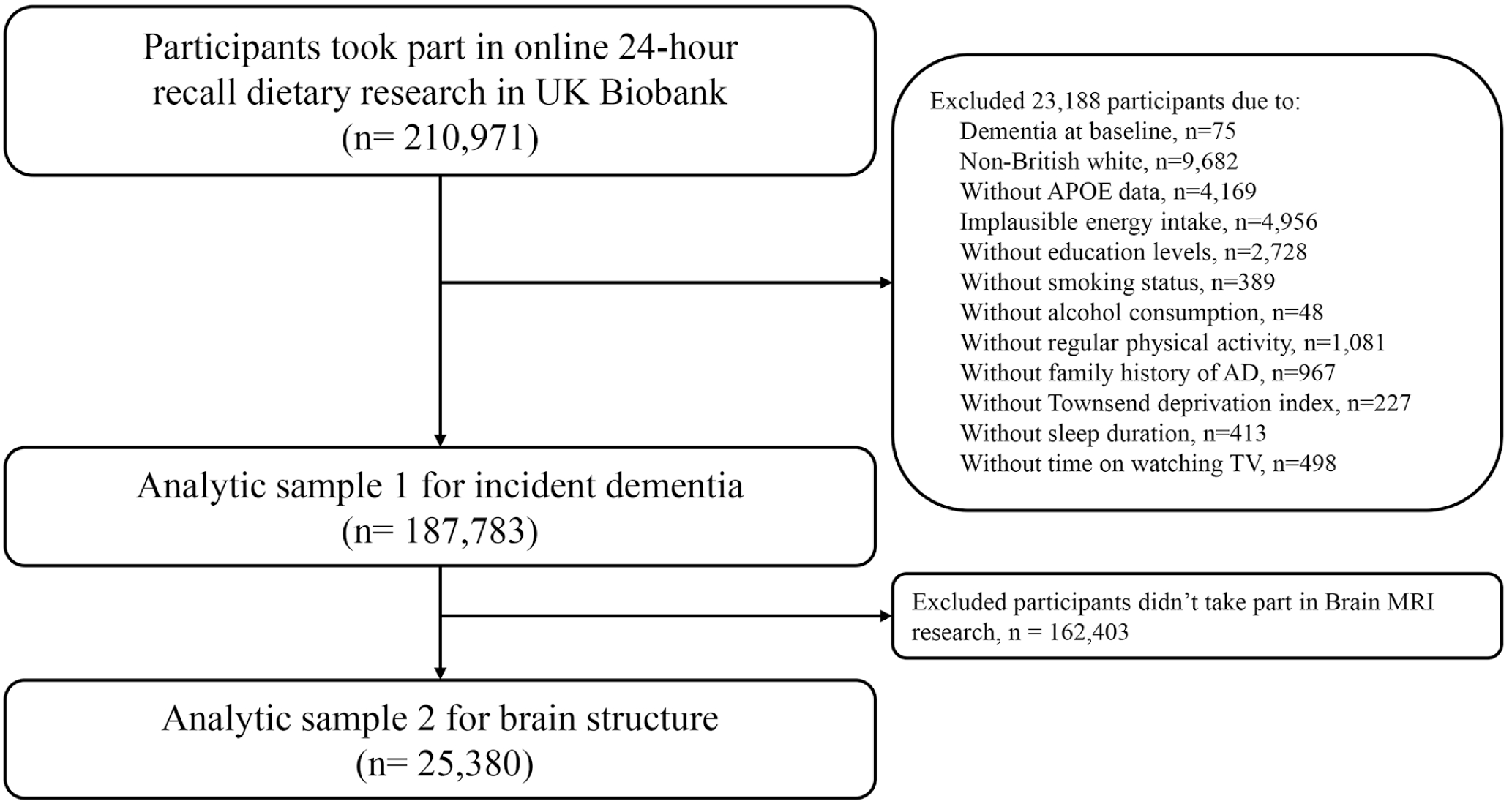
Flow chart of sample selection.

**Figure 3.**
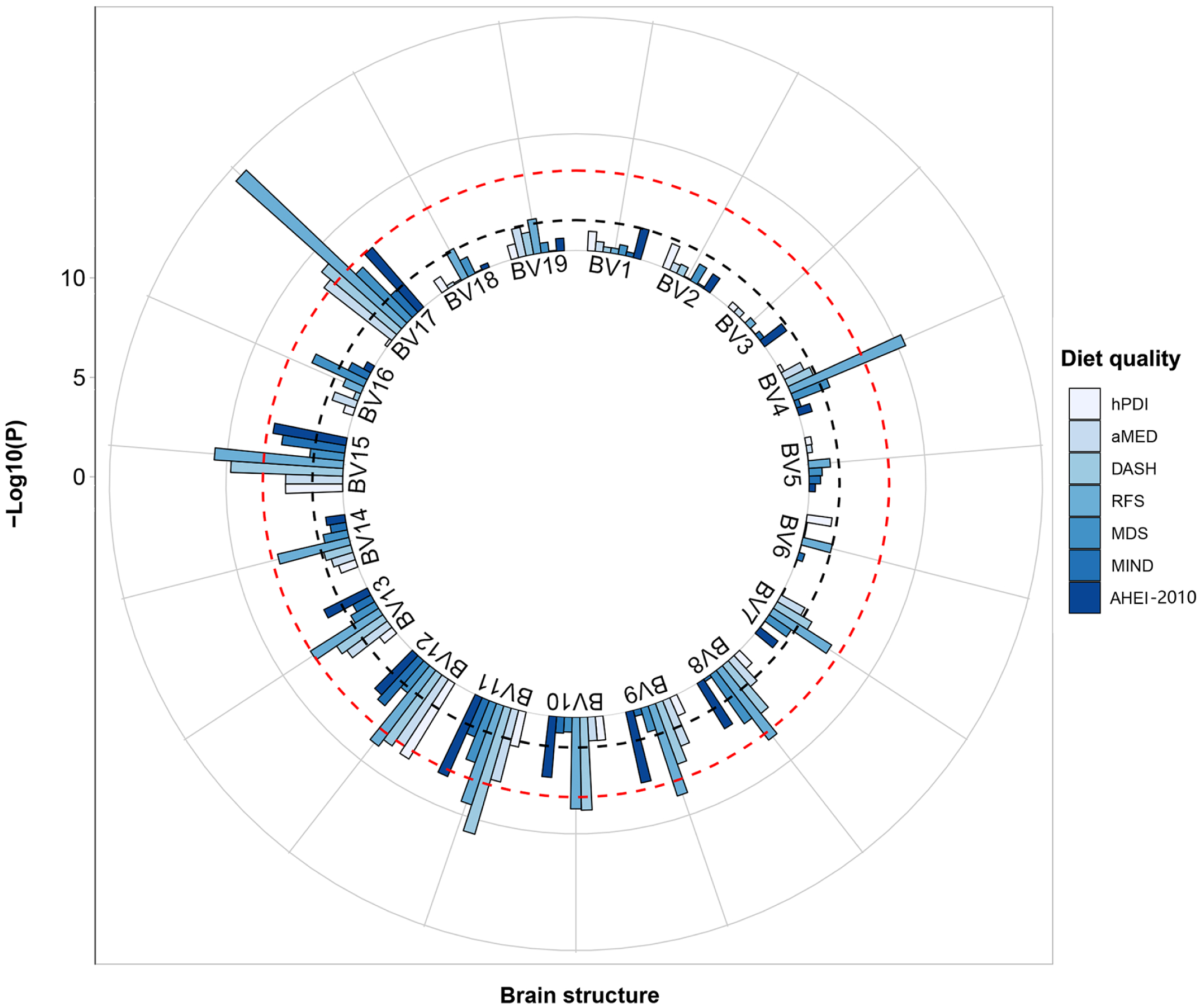
Association between diet quality and brain structures. Notes: hPDI=healthy plant-based diet; MDS= Mediterranean diet score; aMED=alternate Mediterranean diet score; RFS=recommended food score; DASH=dietary approaches to stop hypertension; MIND=Mediterranean-DASH intervention for neurodegenerative delay; AHEI-2010=the alternate healthy eating indicator-2010. BV1-BV19 represents the volumes of 1) total brain, 2) grey matter (GM), 3) white matter (WM), 4) GM in superior frontal gyrus, 5) GM in inferior frontal gyrus, 6) GM in middle frontal gyrus, 7) GM in supplementary motor cortex, 8) GM in precentral gyrus, 9) GM in postcentral gyrus, 10) GM in precuneus, 11) GM in superior parietal lobe, 12) GM in parahippocampal gyrus, 13) GM in middle temporal gyrus, 14) GM in inferior temporal gyrus, 15) hippocampus, 16) putamen, 17) thalamus, 18) caudate, and 19) amygdala. P values were calculated by linear regression based on model 3 and was -log10-transformed. Black dotted line is the significant line of P=0.05, and red dotted line is the significant line after correction for multiple comparisons of P=3.7×10^−4^ (0.05/7/19).

### 2.2 Dietary assessment

UK Biobank assessed more than 200 kinds of foods and 30 kinds of drinks during the previous 24 hours using the Oxford WebQ (https://biobank.ndph.ox.ac.uk/showcase/ukb/docs/DietWebQ.pdf). It was first conducted during the recruitment of the last 70,000 participants. Then, more than 320,000 previous participants with valid e-mail addresses were invited via e-mail. The online questionnaires were completed on four separate occasions over approximately one year (Feb 2011-April 2012).

### 2.3 Diet quality assessment

Seven diet quality scores (i.e., hPDI, MDS, aMED, RFS, DASH, MIND, and AHEI-2010) were considered. As reported in a previous study, there was a strong correlation (i.e., r = 0.88; P < 0.001 for hPDI) between diet quality at baseline (diet quality calculated by dietary assessment at first response to the questionnaire) and average diet quality (diet quality calculated by average dietary assessment of all repeats) in UK Biobank^24^; therefore, dietary assessment of first response was adopted to calculate diet quality scores in this study for simplification. The seven diet quality scores were described briefly below, and details on the components and scoring methods of them are shown in **Supplementary Table S1**.

#### 2.3.1 hPDI

hPDI is an innovative diet quality scores created by Dr. Satija et al^18^ to emphasize intake of healthy plant foods associated with improved health outcomes. The modified hPDI score of the UK Biobank study developed by Heianza et al^24^ consisted of 17 components that categorized into “healthy plant”, “less healthy plant” and “animal” based food. Higher intake of foods from the “healthy plant” food component was given positive scores, while higher intake of foods from the other two food component were given reverse scores. Total scores of these 17 components were summed to obtain the hPDI.

#### 2.3.2 MDS

MDS is a food-based and nutrient-based nine-item score developed and validated by Trichopoulou et al^25^ to reflect adherence to a Mediterranean style diet. It consists of nine components that are scored either 1 or 0 based on sex-specific median intake as cut-offs. Total MDS score ranged from 0 to 9, with higher scores representing better adherence to the Mediterranean diet.

#### 2.3.3 aMED

aMED score developed by Fung et al^26^ was an adaptation of the MDS. It includes nine food components that are common in the Mediterranean-style diet adapted to the US population. The aMED ranged from 0 to 9, with 0 representing the minimum alignment and 9 representing the maximum alignment of the Mediterranean-style diet.

#### 2.3.4 RFS

RFS was developed by Dr. Kant et al^27^ to measure overall diet quality based on consumption of five food components recommended by the 4^th^ edition of the Dietary Guidelines for Americans. Each food component were assigned a score of 1 if consumption above minimum amount threshold (15/g per day for non-beverages and 30/g per day for beverages), or 0 if intakes below this threshold. Total score of RFS ranged from 0 to 21, with higher scores indicating a higher quality of diet and a better consumption of the healthy foods recommended by the dietary guideline.

#### 2.3.5 DASH

The DASH score was created based on foods that were emphasized or discouraged in the DASH trial^28^. It consists of eight food components that are scored 1 to 5 based on quintiles classified according to intake ranking. The modified DASH score developed by previous research within the UK Biobank study^26^ consists of seven components due to lack of sodium intake information. Total score of DASH ranged from 7 to 35, with higher scores indicating better adherence to the DASH diet.

#### 2.3.6 MIND

MIND diet is a combination of the Mediterranean diet and the DASH diet that specifically focuses on brain health developed by Dr. Morris et al^12^. It consists of 10 brain healthy food components and five unhealthy food components. Each food component is scored 1 to 5 based on quintiles classified according to intake ranking. Since olive oil consumption information was not collected in the UK Biobank Study, it was not included in this study. The total MIND score was computed by summing all the 14 components, and ranged from 14 to 70.

#### 2.3.7 AHEI-2010

AHEI-2010 includes foods components that have been found to be associated with risks of major chronic diseases^29^. The original AHEI-2010 score consists of 11 food components that each range from 0 to 10 based on consumption of each food portion. This study included a modified AHEI-2010 consists of 8 components instead due to the lack of information on sodium, trans fat, and long-chain fatty acid intakes in the UK Biobank Study. The total AHEI-2010 score ranged from 0 to 80 in this study.

### 2.4 Outcomes

#### 2.4.1 Primary outcome: incident dementia

The diagnosis of all-cause dementia was obtained by integrating data from two resources: hospital inpatient records and death register data recorded by the International Classification of Diseases (ICD) coding system^22^. The ICD-10 and ICD-9 codes, as well as self-reported data for identifying participants with dementia were listed in **Supplementary Table S2**. The censoring time of incident dementia in our study was Aug 29, 2021. For each participant, time-to-event was calculated as months from the date of first response to the 24-hour recall dietary questionnaire to the date of the first diagnosis of dementia, date of death, date of loss to follow up, or Aug 29, 2021, whichever came first.

#### 2.4.2 Secondary outcome: brain volumes

Brain volumes were extracted from T1-structural brain MRI images, which were provided by an ongoing research that began in 2014, with the aim to acquire high-quality imaging data from 100,000 predominantly healthy participants in UK Biobank^30^. In this study, we used imaging-derived phenotypes (IDPs) according to a previous study, in which the brain MRI imaging processing pipelines were described in detail^31^. In total, 19 IDPs were involved, including volume of grey matter (GM), the volume of white matter (WM), volume of brain (GM+WM), regional grey matter volumes (i.e., volumes of GM in superior frontal gyrus, inferior frontal gyrus, middle frontal gyrus, supplementary motor cortex, precentral gyrus, postcentral gyrus, precuneus, superior parietal lobe, parahippocampal gyrus, middle temporal gyrus, and inferior temporal gyrus), and volumes of several subcortical areas (including hippocampus, putamen, thalamus, caudate, and amygdala). Especially, volumes of grey matter, white matter, and the total brain were normalized by head size^32^. For the other IDPs, the sum of volumes in the left and right hemispheres was calculated.

### 2.5 Covariates

There were several covariates considered in our study, including chronological age, sex, education level, Townsend deprivation index (TDI), body mass index (BMI), smoking status, alcohol consumption, regular physical activity (RPA), time on watching TV, sleep duration, family history of AD, APOE genotype, and cardiovascular disease (CVD), cancer, and diabetes at baseline. In particular, education level was classified as high (college or university degree), intermediate (A/AS levels or equivalent, O levels/GCSEs or equivalent), and low (none of the aforementioned). TDI, an indicator of socioeconomic status, was calculated immediately before a participant joined UK Biobank based on the corresponding area of his/her postcode. BMI was calculated by weight/height^2^ (kg/m^2^) and classified into four categories: underweight (<18.5 kg/m^2^), normal (18.5 to 24.9 kg/m^2^), overweight (25 to 29.9 kg/m^2^), and obese (≥30 kg/m^2^)^33^. Smoking status included never, previous and current smokers. Alcohol consumption was divided into four levels: never or on special occasions only, one to three times per month, one to four times per week, and daily or almost daily. RPA was defined as meeting the current global health recommendations for physical activity (150 minutes of moderate activity or 75 minutes of vigorous activity or an equivalent combination), which equated to ≥500 MET-minutes/week), or no RPA (<500 MET-minutes/week)^34^. APOE genotype was identified based on two single nucleotide polymorphisms (SNPs) including rs7412 and rs429358 (**Supplementary Table S3**), and was classified into E2 (low risk, including ε2/ε2 and ε2/ε3), E3 (neutral risk, ε3/ε3), and E4 (high risk, including ε3/ε4 and ε4/ε4)^35^. In particular, ε2/ε4 was excluded because of its ambiguous genetic risk of dementia^35^.

### 2.6 Statistical analysis

The baseline characteristics of study population were presented by incident dementia. Continuous and categorical variables were described by mean ± standard deviation (SD) and number (percentages) and compared using ANOVA analysis and Chi-square test, respectively.

To estimate the association of diet quality (as tertiles) with incident dementia, Cox proportional hazards regressions were conducted (analysis 1). Three models were applied: model 1 adjusted for chronological age and sex; model 2 further adjusted for education level, TDI, BMI, smoking status, alcohol consumption, RPA, time on watching TV, sleep duration, family history of AD, and APOE genotype; and model 3 additionally adjusted for three chronic diseases at the baseline, including CVD, cancer, and diabetes. The hazard ratios (HRs) and their 95% confidence intervals (CIs) were documented, as well as the P for trend. To estimate the associations of diet quality with brain volumes, linear regression models were conducted (analysis 2). Three same models as above were applied and the coefficient and SD were documented.

To test the robustness of the associations, several sensitivity analyses were conducted. For associations of diet quality with incident dementia and brain volumes (analysis 1 and 2), four sensitivity analyses were performed: 1) to evaluate whether the associations differed by subgroup, we performed stratified analyses by each covariate except for age and chronic diseases; 2) we excluded participants aged less than 60 years as most dementia occur among the older population; 3) we repeated the analyses among relatively healthier participants without CVD, cancer, and diabetes at baseline to minimize the effects of these diseases; 4) we repeated the analyses with additional adjustment for CVD biomarkers including systolic blood pressure (SBP), diastolic blood pressure (DBP), glycosylated hemoglobin (HbA1c), and high-density lipoprotein cholesterol (HDL-C) (model 4). For associations of diet quality with incident dementia (analysis 1), three additional sensitivity analyses were conducted: 1) we repeated the analyses after setting the end of follow-up at the occurrence of COVID-19 (censor date: Nov 30, 2019) to minimize the effects of COVID-19; 2) we repeated the analyses by type of dementia (i.e., Alzheimer’s or vascular dementia); and 3) we repeated the analyses while considering death as a competing risk.

All statistical analyses were conducted by SAS 9.4 (SAS Institute Inc., Cary, NC) and R (version 4.0.3). A two-tailed p < 0.05 was identified as statistically significant.

### 2.7 Patient and Public Involvement statement

Patients and the public were not involved in setting the research question or the outcome measures, and developing plans for recruitment, design, or implementation of the study. No plans exist to involve patients in dissemination.

## 3 Results

### 3.1 Basic characteristics

The analytic sample 1 included 187,783 participants, and the mean age was 56.8 years, with 54.9% females. The total follow-up of analytic sample 1 was 1,969,993 person-years (average 10.5 years per person). About 0.73% (1,363/187,783) participants developed dementia during the follow-up. Participants with incident dementia during the follow-up were more likely to be older, males, obese, previous smokers, and have lower education levels, have higher alcohol consumption or never drinking, have longer sleep durations, have longer time on watching TV, have family histories of AD, and have chronic diseases at baseline (all P<0.05, **Table 1**). The characteristics of analytic sample 2 are exhibited in **Supplementary Table S4**.

**Table 1:** Basic characteristics of 187,783 participants from UK Biobank.

|  | **Total** | **Non-incident dementia<sup>a</sup>** | **Incident Dementia<sup>a</sup>** | **P<sup>b</sup>** |
| --- | --- | --- | --- | --- |
|  | **N=187,783** | **N=186,420** | **N=1,363** |  |
| **Age** | 56.8 (7.88) | 56.7 (7.88) | 64.2 (4.88) | **<0.001** |
| **Sex** |  |  |  | **<0.001** |
| Female | 103,054 (54.9%) | 102,446 (55.0%) | 608 (44.6%) |  |
| Male | 84,729 (45.1%) | 83,974 (45.0%) | 755 (55.4%) |  |
| **Education levels<sup>c</sup>** |  |  |  | **<0.001** |
| High | 80,911 (43.1%) | 80,455 (43.2%) | 456 (33.5%) |  |
| Intermediate | 63,535 (33.8%) | 63,069 (33.8%) | 466 (34.2%) |  |
| Low | 43,337 (23.1%) | 42,896 (23.0%) | 441 (32.4%) |  |
| **Townsend deprivation index** | -1.70<br>(2.79) | -1.70 (2.79) | -1.67 (2.87) | 0.642 |
| **BMI (kg/m<sup>2</sup>)<sup>d</sup>** |  |  |  | **<0.001** |
| Underweight | 1004 (0.54%) | 994 (0.53%) | 10 (0.74%) |  |
| Normal | 69,770 (37.2%) | 69,313 (37.3%) | 457 (33.7%) |  |
| Overweight | 78,092 (41.7%) | 77,554 (41.7%) | 538 (39.7%) |  |
| Obese | 38,548 (20.6%) | 38,197 (20.5%) | 351 (25.9%) |  |
| **Smoking status** |  |  |  | **<0.001** |
| Never | 105,586 (56.2%) | 104,936 (56.3%) | 650 (47.7%) |  |
| Previous | 67,842 (36.1%) | 67,226 (36.1%) | 616 (45.2%) |  |
| Current | 14,355 (7.64%) | 14,258 (7.65%) | 97 (7.12%) |  |
| **Alcohol consumption<sup>e</sup>** |  |  |  | **<0.001** |
| None | 27,152 (14.5%) | 26,877 (14.4%) | 275 (20.2%) |  |
| Low | 20,378 (10.9%) | 20,220 (10.8%) | 158 (11.6%) |  |
| Intermediate | 95,753 (51.0%) | 95,173 (51.1%) | 580 (42.6%) |  |
| High | 44,500 (23.7%) | 44,150 (23.7%) | 350 (25.7%) |  |
| **Regular physical activity<sup>f</sup>** |  |  |  | 0.086 |
| No | 81,451 (43.4%) | 80,828 (43.4%) | 623 (45.7%) |  |
| Yes | 106,332 (56.6%) | 105,592 (56.6%) | 740 (54.3%) |  |
| **Sleep duration (hour)** | 7.17 (0.99) | 7.17 (0.99) | 7.27 (1.21) | **0.002** |
| **Time on watching TV (hour)** | 1.87 (3.36) | 1.87 (3.36) | 2.64 (2.93) | **<0.001** |
| **Family history of Alzheimer's disease** |  |  |  | **<0.001** |
| No | 162,372 (86.5%) | 161,360 (86.6%) | 1,012 (74.2%) |  |
| Yes | 25,411 (13.5%) | 25,060 (13.4%) | 351 (25.8%) |  |
| **Primary diseases at baseline** |  |  |  |  |
| **Cancer** |  |  |  | **<0.001** |
| No | 171,651 (91.4%) | 170,444 (91.4%) | 1,207 (88.6%) |  |
| Yes | 16,132 (8.59%) | 15,976 (8.57%) | 156 (11.4%) |  |
| **CVD** |  |  |  | **<0.001** |
| No | 176,987 (94.3%) | 175,869 (94.3%) | 1,118 (82.0%) |  |
| Yes | 10,796 (5.75%) | 10,551 (5.66%) | 245 (18.0%) |  |
| **Diabetes** |  |  |  | **<0.001** |

|  | Total | Non-incident<br>dementia<sup>a</sup> | Incident<br>Dementia<sup>a</sup> | P<sup>b</sup> |
| --- | --- | --- | --- | --- |
|  | N=187,783 | N=186,420 | N=1,363 |  |
| No | 185,522 (98.8%) | 184,220 (98.8%) | 1,302 (95.5%) |  |
| Yes | 2,261 (1.20%) | 2,200 (1.18%) | 61 (4.48%) |  |
| **Diet quality assessment (continuous)** |  |  |  |  |
| hPDI | 54.0 (6.96) | 54.0 (6.97) | 54.5 (6.65) | **0.003** |
| MDS | 4.02 (1.56) | 4.02 (1.56) | 3.88 (1.58) | **0.001** |
| aMED | 3.70 (1.80) | 3.70 (1.80) | 3.56 (1.81) | **0.003** |
| RFS | 10.3 (4.00) | 10.3 (4.00) | 9.91 (4.02) | **0.001** |
| DASH | 20.2 (4.56) | 20.2 (4.56) | 20.1 (4.34) | 0.176 |
| MIND | 37.1 (7.12) | 37.1 (7.12) | 36.9 (7.05) | 0.134 |
| AHEI-2010 | 37.4 (12.2) | 37.4 (12.2) | 37.0 (11.8) | 0.231 |
| **Diet quality assessment (categorical)** |  |  |  |  |
| hPDI |  |  |  | **0.003** |
| Tertile 1 | 66,459 (35.4%) | 66,029 (35.4%) | 430 (31.5%) |  |
| Tertile 2 | 63,339 (33.7%) | 62,827 (33.7%) | 512 (37.6%) |  |
| Tertile 3 | 57,985 (30.9%) | 57,564 (30.9%) | 421 (30.9%) |  |
| MDS |  |  |  |  |
| Tertile 1 | 71,396 (38.0%) | 70,796 (38.0%) | 600 (44.0%) |  |
| Tertile 2 | 83,239 (44.3%) | 82,693 (44.4%) | 546 (40.1%) |  |
| Tertile 3 | 33,147 (17.7%) | 32,930 (17.7%) | 217 (15.9%) |  |
| aMED |  |  |  | **0.010** |
| Tertile 1 | 89,316 (47.6%) | 88,613 (47.5%) | 703 (51.6%) |  |
| Tertile 2 | 65,835 (35.1%) | 65,387 (35.1%) | 448 (32.9%) |  |
| Tertile 3 | 32,630 (17.4%) | 32,418 (17.4%) | 212 (15.6%) |  |
| RFS |  |  |  | **0.011** |
| Tertile 1 | 64,024 (34.1%) | 63,518 (34.1%) | 506 (37.1%) |  |
| Tertile 2 | 63,789 (34.0%) | 63,320 (34.0%) | 469 (34.4%) |  |
| Tertile 3 | 59,970 (31.9%) | 59,582 (32.0%) | 388 (28.5%) |  |
| DASH |  |  |  | 0.184 |
| Tertile 1 | 70,189 (37.4%) | 69,648 (37.4%) | 541 (39.7%) |  |
| Tertile 2 | 59,777 (31.8%) | 59,352 (31.8%) | 425 (31.2%) |  |
| Tertile 3 | 57,817 (30.8%) | 57,420 (30.8%) | 397 (29.1%) |  |
| MIND |  |  |  | 0.077 |
| Tertile 1 | 71,574 (38.1%) | 71,014 (38.1%) | 560 (41.1%) |  |
| Tertile 2 | 58,821 (31.3%) | 58,415 (31.3%) | 406 (29.8%) |  |
| Tertile 3 | 57,388 (30.6%) | 56,991 (30.6%) | 397 (29.1%) |  |
| AHEI-2010 |  |  |  | 0.064 |
| Tertile 1 | 63,988 (34.1%) | 63,488 (34.1%) | 500 (36.7%) |  |
| Tertile 2 | 61,211 (32.6%) | 60,767 (32.6%) | 444 (32.6%) |  |
| Tertile 3 | 62,584 (33.3%) | 62,165 (33.3%) | 419 (30.7%) |  |
| **APOE genotype<sup>g</sup>** |  |  |  | **<0.001** |
| E2 | 24,137 (12.9%) | 24,043 (12.9%) | 94 (6.90%) |  |
| E3 | 110,706 (59.0%) | 110,168 (59.1%) | 538 (39.5%) |  |
| E4 | 52,940 (28.2%) | 52,209 (28.0%) | 731 (53.6%) |  |

Notes: the continuous and categorical variables were described by mean (standard deviation) and number (proportion), respectively. BMI=body mass index; CVD= cardiovascular disease; hPDI=healthy plant-based diet; MDS= Mediterranean diet score; aMED=alternate Mediterranean diet score; RFS=recommended food score; DASH=dietary approaches to stop hypertension; MIND=Mediterranean-DASH intervention for neurodegenerative delay; AHEI-2010=the alternate healthy eating indicator-2010.
<sup>a</sup>The censor date of incident dementia is Aug 29, 2021, and the baseline was defined as the date of the first time that a participant completed online 24-hour recall dietary questionnaire.
<sup>b</sup>Chi-square test and ANOVA analysis were used for comparison of continuous and categorical variables, respectively.
<sup>c</sup>Education levels were defined as high (college or university degree), intermediate (A/AS levels or equivalent, O levels/GCSEs or equivalent), and low (none of the aforementioned).
<sup>d</sup>BMI was calculated by the body mass divided by the square of the body height. It was classified into four categories: underweight (under 18.5 kg/m<sup>2</sup>), normal (18.5 to 24.9 kg/m<sup>2</sup>), overweight (25 to 29.9 kg/m<sup>2</sup>) and obese (over 30 kg/m<sup>2</sup>).
<sup>e</sup>Alcohol consumption was defined as none (never or special occasions only), low(one to three times per month), intermediate (one to four times per week), or high (daily or almost daily).
<sup>f</sup>Regular physical activity was defined as meeting the current global health recommendations for physical activity (150 minutes of moderate activity or 75 minutes of vigorous activity or an equivalent combination), which equated to $\geq 500$ MET-minutes/week, or no regular physical activity (<500 MET-minutes/ week).
<sup>g</sup>APOE genotypes were defined as E2 (protective), E3 (mild), or E4 (risky) by two single-nucleotide polymorphisms, rs429358 and rs7412, the details were described in supplementary table S1.

### 3.2 Associations between midlife diet quality and incident dementia

As shown in **Table 2**, after adjusting for chronological age and sex (model 1), higher diet quality scores (except for hPDI) were associated with a lower incidence risk of dementia (all P for trend<0.001). For instance, for RFS, the HRs of the intermediate tertile group (Tertile 2) and the highest tertile group (Tertile 3) relative to the lowest tertile group (Tertile 1) were 0.82 (95%CI=0.72 to 0.93) and 0.65 (95%CI=0.57 to 0.74), respectively. Moreover, higher RFS was linearly related with lower risk of incident dementia, with P for trend<0.001. In addition, the directions and magnitudes of these associations did not change substantially after adjusting for more covariables in model 2 (further adjusted for education level, TDI, BMI, smoking status, alcohol consumption, RPA, time on watching TV, sleep duration, family history of AD, and APOE genotype) and model 3 (additionally adjusted for three chronic diseases at the baseline, including CVD, cancer, and diabetes).

**Table 2.** Associations of diet quality with incident dementia in UK Biobank.

| Diet quality |  | Model 1<sup>a</sup> |  |  | Model 2<sup>b</sup> |  |  | Model 3<sup>c</sup> |  |  |
| --- | --- | --- | --- | --- | --- | --- | --- | --- | --- | --- |
|  |  | HR | 95% CI | P | HR | 95% CI | P | HR | 95% CI | P |
| **hPDI** | Tertile 1 (ref) |  |  |  |  |  |  |  |  |  |
|  | Tertile 2 | 1.12 | (0.98, 1.27) | 0.085 | 1.13 | (0.99, 1.29) | 0.065 | 1.12 | (0.98, 1.27) | 0.085 |
|  | Tertile 3 | 1.02 | (0.89, 1.17) | 0.768 | 1.03 | (0.90, 1.18) | 0.672 | 1.02 | (0.89, 1.17) | 0.747 |
|  | P for trend |  | 0.754 |  |  | 0.654 |  |  | 0.731 |  |
| **MDS** | Tertile 1 (ref) |  |  |  |  |  |  |  |  |  |
|  | Tertile 2 | 0.74 | (0.66, 0.83) | <0.001 | 0.77 | (0.69, 0.87) | <0.001 | 0.77 | (0.69, 0.87) | <0.001 |
|  | Tertile 3 | 0.71 | (0.60, 0.82) | <0.001 | 0.76 | (0.65, 0.89) | <0.001 | 0.75 | (0.64, 0.88) | <0.001 |
|  | P for trend |  | <0.001 |  |  | <0.001 |  |  | <0.001 |  |
| **aMED** | Tertile 1 (ref) |  |  |  |  |  |  |  |  |  |
|  | Tertile 2 | 0.80 | (0.71, 0.90) | <0.001 | 0.84 | (0.74, 0.94) | 0.003 | 0.83 | (0.74, 0.94) | 0.003 |
|  | Tertile 3 | 0.72 | (0.62, 0.84) | <0.001 | 0.77 | (0.66, 0.91) | 0.002 | 0.77 | (0.66, 0.91) | 0.002 |
|  | P for trend |  | <0.001 |  |  | <0.001 |  |  | <0.001 |  |
| **RFS** | Tertile 1 (ref) |  |  |  |  |  |  |  |  |  |
|  | Tertile 2 | 0.82 | (0.72, 0.93) | 0.002 | 0.85 | (0.75, 0.97) | 0.014 | 0.85 | (0.75, 0.97) | 0.014 |
|  | Tertile 3 | 0.65 | (0.57, 0.74) | <0.001 | 0.70 | (0.61, 0.80) | <0.001 | 0.70 | (0.61, 0.81) | <0.001 |
|  | P for trend |  | <0.001 |  |  | <0.001 |  |  | <0.001 |  |
| **DASH** | Tertile 1 (ref) |  |  |  |  |  |  |  |  |  |
|  | Tertile 2 | 0.78 | (0.69, 0.89) | <0.001 | 0.81 | (0.71, 0.92) | 0.001 | 0.81 | (0.71, 0.92) | 0.002 |
|  | Tertile 3 | 0.73 | (0.64, 0.83) | <0.001 | 0.76 | (0.67, 0.87) | <0.001 | 0.77 | (0.67, 0.88) | <0.001 |
|  | P for trend |  | <0.001 |  |  | <0.001 |  |  | <0.001 |  |
| **MIND** | Tertile 1 (ref) |  |  |  |  |  |  |  |  |  |
|  | Tertile 2 | 0.84 | (0.74, 0.95) | 0.007 | 0.88 | (0.77, 1.00) | 0.045 | 0.88 | (0.77, 1) | 0.049 |
|  | Tertile 3 | 0.80 | (0.71, 0.92) | 0.001 | 0.87 | (0.76, 1.00) | 0.045 | 0.87 | (0.76, 0.99) | 0.040 |
|  | P for trend |  | 0.001 |  |  | 0.037 |  |  | 0.033 |  |
| **AHEI-2010** | Tertile 1 (ref) |  |  |  |  |  |  |  |  |  |
|  | Tertile 2 | 0.80 | (0.70, 0.91) | 0.001 | 0.83 | (0.73, 0.95) | 0.006 | 0.84 | (0.74, 0.95) | 0.008 |
|  | Tertile 3 | 0.70 | (0.61, 0.80) | <0.001 | 0.75 | (0.66, 0.86) | <0.001 | 0.75 | (0.66, 0.86) | <0.001 |
|  | P for trend |  | <0.001 |  |  | <0.001 |  |  | <0.001 |  |

Notes: OR=odds ratio; CI=confident interval; hPDI=healthy plant-based diet; MDS= Mediterranean diet score; aMED=alternate Mediterranean diet score; RFS=recommended food score; DASH=dietary approaches to stop hypertension; MIND=Mediterranean-DASH intervention for neurodegenerative delay; AHEI-2010=the alternate healthy eating indicator-2010. The higher tertiles indicate better diet quality.
<sup>a</sup>Model 1 adjusted for age and sex;
<sup>b</sup>Model 2 further adjusted for education levels, Townsend deprivation index, body mass index, smoking status, alcohol consumption, regular physical activity, sleep duration, time on watching TV, family history of Alzheimer's disease, and APOE genotypes.
<sup>c</sup>Model 3 further adjusted for primary diseases at the time of 24-hour dietary recall research, i.e., cancer, cardiovascular disease, and diabetes.

### 3.3 Associations between midlife diet quality and brain volumes

As shown in **Supplementary Table S5**, after adjusting for all covariables, higher diet quality scores were significantly associated with larger brain volumes with positive coefficients of linear regressions for most IDPs except for volumes of GM, WM, and total brain (GM+WM). Most diet quality scores were positively associated with volumes of GM in the parietal and temporal cortex and volumes of hippocampus and thalamus. Among all diet quality scores, RFS showed the strongest associations with almost all regional grey matter volumes and volumes of subcortical areas. For instance, higher RFS was associated with larger volumes of GM in the postcentral gyrus (β=16.05±4.08, P<0.001, model 3) and the hippocampus (β=5.87±1.26, P<0.001, model 3). However, hPDI and MIND were associated with only three or four IDPs in model 3.

### 3.4 Sensitivity analyses of the associations between midlife diet quality and incident dementia

For subgroup analyses (**Supplementary Table S6**), the associations between most diet quality scores (i.e., MDS, aMED, DASH, RFS, and AHEI-2010) and incident dementia were relatively robust by subgroup, except that the associations between MIND and incident dementia were attenuated in several subgroups. For the additional six sensitivity analyses (**Supplementary Table S7**), we found that the main results were maintained after excluding participants below 60 years old (Sensitivity analysis 1), excluding participants with chronic disease (Sensitivity analysis 2), setting the end of follow-up at the occurrence of COVID-19 pandemic (Sensitivity analysis 4), or running competing risk model (Sensitivity analysis 6). However, MIND was not associated with incident dementia after further adjusting for some CVD biomarkers (Sensitivity analysis 3), and the associations between all diet quality scores (i.e., MDS, aMED, DASH, RFS, MIND, and AHEI-2010) and incident dementia were attenuated in stratified analysis by type of dementia (AD vs. vascular, Sensitivity analysis 5.1 and 5.2).

### 3.5 Sensitivity analyses of the associations between midlife diet quality and brain volumes

For subgroup analyses (**Supplementary Table S8**), the patterns of associations between diet quality and brain volumes were maintained, and RFS was still associated with most IDPs across subgroups. For the additional three sensitivity analyses (**Supplementary Table S9**), we found that the associations between diet quality and brain volumes were maintained when excluding participants with chronic diseases at baseline (Sensitivity analysis 2) or further adjusting for CVD biomarkers (Sensitivity analysis 3). However, after excluding participants less than 60 years old (Sensitivity analysis 1), AHEI-2010 became positively associated with larger volumes of brain and WM after adjusting for all covariates.

## 4. Discussion

Based on a large sample of over 180,000 middle-aged and older adults from UK

, we found that six diet quality scores (i.e., MDS, aMED, RFS, DASH, MIND, and AHEI-2010) but not hPDI were significantly associated with the higher risk of incident dementia after an average follow-up of 10.5 years. Furthermore, all of the diet quality scores were positively associated with larger brain volumes in specific regions, particularly the parietal and temporal cortex, as well as the hippocampus in over 25,000 adults. This study shows a comprehensive picture of the consistent associations of diet quality with dementia risk and brain health, providing further evidence and mechanistic insights into the role of diet quality in slowing down the progression of dementia.

The beneficial effect of healthy diets on cognitive function has previously been explored^36 37^. However, limited research has estimated the associations between diet quality and the risk of incident dementia^11 12 16 17 38-45^. Most studies focused on MDS^16 17 39-41^, with a few studies considering aMDS^11 45^, DASH^11 12 17 42^, MIND^12 42-44^, and AHEI-2010^11^. Moreover, the results of these studies were inconsistent. Significant associations of MDS^39-41^, DASH^12^, and MIND^12 43 44^ with reduced dementia risk were observed in short follow-up time (i.e., 3 to 7 years) studies. However, the associations diminished with longer follow-up time (i.e., 12 to 27 years)^11 17 38 45^. After a median follow-up time (i.e., 10.5 years), we observed significant associations of diet quality with the risk of incident dementia in the current study. The inconsistency across studies may be partly explained by the differences in follow-up time and study population. Dietary habits may be affected by the cognitive decline during the long preclinical period before dementia^46^; and thus, reverse causation may exist in these short-term follow-up studies. Additionally, previous studies only assessed one diet quality score^16 40^ or a few diet quality scores simultaneously^11 12 17^, thus limiting the comparability across diet quality scores. In contrast, we systematically assessed diet quality with various diet quality scores in the same large sample of UK middle-aged and older adults. Although most diet quality scoring systems showed that higher quality diets were protective of incident dementia while hPDI was not associated. A potential explanation might be that when more food items were considered in hPDI (188 kinds of food in this study), the influence of some critical food on dementia was attentued^24^. Moreover, the absence of data on vegetable oil to construct hPDI in UK Biobank may have influenced our results, as vegetable oil is a major source of dietary fatty acids, which could help maintain cognitive functioning^14^. In addition, the results raised an intriguing question of whether the hPDI is suitable for the UK population. More studies are required to assess the true effect of hPDI on dementia.

Since the neuropathologic changes of dementia may begin 20 years before dementia diagnosis^47^, exploring the brain structures may help understand potential mechanisms that relate diet quality to dementia risk. Accordingly, we found that all considered diet quality scores were associated with larger volumes of several specific regions (e.g., parietal and temporal cortex, and hippocampus), but not with total volumes of the total brain (i.e., GM and WM), GM, and WM. To date, limited studies have explored the associations of diet quality scores including MDS^21 48-50^, RFS^50^, MIND^44 51 52^, and AHEI-2010^53^ with brain structures, mainly among older adults with a relatively small sample size^21 44 48 49 54^, and reported mixed results. With over 25,000 middle-aged and older adults, this study revealed discrepancies in various diet quality scores. RFS presented to be the most sensitive diet quality scores to brain volumes, with significant associations with almost all regional grey matter volumes and volumes of subcortical areas. However, hPDI and MIND were the least sensitive diet quality scores to brain volumes. These results are in line with differences in the magnitude of the associations between diet quality scores and the risk of incident dementia, confirming the differences in various diet quality scores and providing evidence for the necessity of comprehensive evaluation of diet quality.

We speculate that a potential mechanism for the benefit of healthy diets in preventing dementia is that adhering to healthy diets may promote brain volumes. Although one or more diet quality scores are significantly associated with almost all regional brain volumes in our study, the most extensively influenced regions include the parietal and temporal cortex, and hippocampus. Similarly, these brain regions are also found to be largely affected by alcohol intake^55 56^, which is an item in the MDS, aMED and AHEI-2010 diets. Volume loss in the temporal lobe and hippocampus have been acknowledged as predictive biomarkers of incident dementia^57^. More recently, the role of the parietal lobe in the development of dementia has attracted attention^58^. The close connectivity between the parietal lobe and other brain areas, as well as a wide range of cognitive function that relied on the parietal lobe, may account for the involvement of the parietal lobe in dementia^58^. Indeed, the development of dementia should be a consequence of changes in multiple brain regions. Nevertheless, due to the cross-sectional design for the second aim of our study, we were unable to draw a temporal association. More longitudinal and experimental studies are needed to verify the underlying mechanisms.

## Strengths and limitations

Major strengths of this study include the larger sample size, the prospective design with long-term follow-up, available APOE genotypes, seven considered diet quality scores, diagnosis of incident dementia through linked hospital data, and assessment of brain structure in a subgroup, providing a comprehensive exhibition of associations of different diet quality scores with incident dementia and brain volumes in the same population, a valuable data resource for future researchers. Other strength includes the series of sensitivity analyses to reinforce our findings. Nevertheless, this study has a few limitations. First, the incidence of dementia in our study was lower than that in other cohort studies^59^, and this may be explained by the fact that UK Biobank participants are healthier than the general UK population. However, such ascertainment of dementia cases has been demonstrated to be in agreement with primary care records^60^. Second, when constructing these diet quality scores, some food items were unavailable in UK Biobank. For example, total sodium intake, as an important food component for assessing the DASH diet, was not available in the UK Biobank dataset. Thus, the diet quality scores we constructed may not be able to fully reflect the diet quality scores per se. Third, we used a single 24-hour dietary recall to assess diet quality, and thereby, recall bias and a lack of representation of habitual diet quality were inevitable. However, previous studies in the UK Biobank showed good correlations of these scores during a period^24^. Finally, although we estimated the associations between diet quality and brain volumes assessed by MRI, the cross-sectional design hampered the ability to explore underlying mechanisms. Meanwhile, the limited events of incident dementia (42/25380, 0.17%) among participants with MRI data led to the difficulty in estimating potential effects of brain volumes in the association between diet quality and incident dementia.

## Conclusion

In this large sample of UK Biobank, we demonstrated that greater adherence to MDS, aMED, RFS, DASH, MIND, and AHEI-2010 at midlife were independently associated with lower risk of incident dementia and larger brain volumes in specific regions, regardless of social-economic status and APOE genotypes. Our study presents a full picture of the potential comprehensive beneficial effect of midlife healthy diets on dementia risk and brain health. These findings underscore the importance of midlife diet quality in maintaining brain health and provide mechanistic insight into the role of diet quality in the prevention of dementia. From a public health perspective, preventive and interventional dietary strategies could counter the growing burden of dementia in aging populations, which may be effective even in resource-limited settings.

## Data Availability

All data produced are available online at http://www.ukbiobank.ac.uk/register-apply.

## Acknowledgments

This study was based on the Application 61856 in UK Biobank. We acknowledge the UK Biobank participants who provided the sample that made the data available.

## Ethical approval

The UK Biobank study was approved by the North West Multi-Centre Research Ethics Committee (11/NW/0382) and written informed consent was provided by each participant before the study. UK Biobank also has approval from the North West Multi-Centre Research Ethics Committee as a Research Tissue Bank (RTB) approval in 2011 and is renewed every 5 years. According to RTB approval, researchers are allowed to use data from UK Biobank without an additional ethical clearance.

## Author contributors

ZL contributed to the conception and design of the work. JZ performed the analysis. JZ, XC, XL, XS, GY, MH, CS, and ZL contributed to the interpretation of data. JZ, XC, and XL wrote the initial draft of the manuscript. XL, XS, GY, MH, CS, YX, HH, TSHJ, GOA, WL, XZ, XG, HA, and ZL revised it critically for important intellectual content. All authors approved the final version of the manuscript. ZL is the guarantor. The corresponding author attests that all listed authors meet authorship criteria and that no others meeting the criteria have been omitted.

## Data sharing

This study was based on the Application 61856 in UK Biobank.

## Transparency statement

The lead author (ZL) affirms that the manuscript is an honest, accurate, and transparent account of the study being reported; that no important aspects of the study have been omitted; and that any discrepancies from the study as originally planned have been explained.

## Dissemination to participants and related patient and public communities

The results of the research will be disseminated to the public through broadcasts and popular science articles.

## References

1. Dementia: The World Health Organization September 2021.

2. Fillit H, Nash DT, Rundek T, et al. Cardiovascular risk factors and dementia. Am J Geriatr Pharmacother 2008;6(2):100–18. doi: 10.1016/j.amjopharm.2008.06.004

3. Cerejeira J, Lagarto L, Mukaetova-Ladinska EB. Behavioral and psychological symptoms of dementia. Front Neurol 2012;3:73. doi: 10.3389/fneur.2012.00073 [published Online First: 20120507]

4. Witthaus E, Ott A, Barendregt JJ, et al. Burden of mortality and morbidity from dementia. Alzheimer Dis Assoc Disord 1999;13(3):176–81. doi: 10.1097/00002093-199907000-00011

5. Livingston G, Huntley J, Sommerlad A, et al. Dementia prevention, intervention, and care: 2020 report of the Lancet Commission. Lancet 2020;396(10248):413–46. doi: 10.1016/s0140-6736(20)30367-6 [published Online First: 20200730]

6. Garcia-Ptacek S, Farahmand B, Kåreholt I, et al. Mortality risk after dementia diagnosis by dementia type and underlying factors: a cohort of 15,209 patients based on the Swedish Dementia Registry. J Alzheimers Dis 2014;41(2):467–77. doi: 10.3233/jad-131856

7. Banerjee S, Smith SC, Lamping DL, et al. Quality of life in dementia: more than just cognition. An analysis of associations with quality of life in dementia. J Neurol Neurosurg Psychiatry 2006;77(2):146–8. doi: 10.1136/jnnp.2005.072983

8. Alfakhri AS, Alshudukhi AW, Alqahtani AA, et al. Depression Among Caregivers of Patients With Dementia. Inquiry 2018;55:46958017750432. doi: 10.1177/0046958017750432

9. Coen RF, O’Boyle CA, Coakley D, et al. Individual quality of life factors distinguishing low-burden and high-burden caregivers of dementia patients. Dement Geriatr Cogn Disord 2002;13(3):164–70. doi: 10.1159/000048648

10. Liu C, Fabius CD, Howard VJ, et al. Change in Social Engagement among Incident Caregivers and Controls: Findings from the Caregiving Transitions Study. J Aging Health 2021;33(1-2):114–24. doi: 10.1177/0898264320961946 [published Online First: 20200923]

11. Hu EA, Wu A, Dearborn JL, et al. Adherence to Dietary Patterns and Risk of Incident Dementia: Findings from the Atherosclerosis Risk in Communities Study. J Alzheimers Dis 2020;78(2):827–35. doi: 10.3233/JAD-200392 [published Online First: 2020/10/13]

12. Morris MC, Tangney CC, Wang Y, et al. MIND diet associated with reduced incidence of Alzheimer’s disease. Alzheimers Dement 2015;11(9):1007–14. doi: 10.1016/j.jalz.2014.11.009 [published Online First: 2015/02/15]

13. Athanasopoulos D, Karagiannis G, Tsolaki M. Recent Findings in Alzheimer Disease and Nutrition Focusing on Epigenetics. Adv Nutr 2016;7(5):917–27. doi: 10.3945/an.116.012229 [published Online First: 20160915]

14. Solfrizzi V, Frisardi V, Capurso C, et al. Dietary fatty acids in dementia and predementia syndromes: epidemiological evidence and possible underlying mechanisms. Ageing Res Rev 2010;9(2):184–99. doi: 10.1016/j.arr.2009.07.005 [published Online First: 2009/08/01]

15. Pelletier A, Barul C, Féart C, et al. Mediterranean diet and preserved brain structural connectivity in older subjects. Alzheimers Dement 2015;11(9):1023–31. doi: 10.1016/j.jalz.2015.06.1888 [published Online First: 20150717]

16. Féart C, Samieri C, Rondeau V, et al. Adherence to a Mediterranean diet, cognitive decline, and risk of dementia. JAMA 2009;302(6):638–48.

17. Larsson SC, Wolk A. The Role of Lifestyle Factors and Sleep Duration for Late-Onset Dementia: A Cohort Study. J Alzheimers Dis 2018;66(2):579–86.

18. Satija A, Bhupathiraju SN, Spiegelman D, et al. Healthful and Unhealthful Plant-Based Diets and the Risk of Coronary Heart Disease in U.S. Adults. J Am Coll Cardiol 2017;70(4):411–22. doi: 10.1016/j.jacc.2017.05.047 [published Online First: 2017/07/22]

19. Apostolova LG, Thompson PM. Mapping progressive brain structural changes in early Alzheimer’s disease and mild cognitive impairment. Neuropsychologia 2008;46(6):1597–612. doi: 10.1016/j.neuropsychologia.2007.10.026 [published Online First: 20071214]

20. Titova OE, Ax E, Brooks SJ, et al. Mediterranean diet habits in older individuals: associations with cognitive functioning and brain volumes. Exp Gerontol 2013;48(12):1443–8. doi: 10.1016/j.exger.2013.10.002 [published Online First: 20131011]

21. Gu Y, Brickman AM, Stern Y, et al. Mediterranean diet and brain structure in a multiethnic elderly cohort. Neurology 2015;85(20):1744–51. doi: 10.1212/WNL.0000000000002121 [published Online First: 2015/10/23]

22. Lourida I, Hannon E, Littlejohns TJ, et al. Association of Lifestyle and Genetic Risk With Incidence of Dementia. Jama 2019;322(5):430–37. doi: 10.1001/jama.2019.9879 [published Online First: 2019/07/16]

23. Merino J, Dashti HS, Sarnowski C, et al. Genetic analysis of dietary intake identifies new loci and functional links with metabolic traits. Nature Human Behaviour 2022;6(1):155–63. doi: 10.1038/s41562-021-01182-w

24. Heianza Y, Zhou T, Sun D, et al. Healthful plant-based dietary patterns, genetic risk of obesity, and cardiovascular risk in the UK biobank study. Clinical Nutrition 2021;40(7):4694–701. doi: https://doi.org/10.1016/j.clnu.2021.06.018

25. Trichopoulou A, Orfanos P, Norat T, et al. Modified Mediterranean diet and survival: EPIC-elderly prospective cohort study. Bmj 2005;330(7498):991. doi: 10.1136/bmj.38415.644155.8F [published Online First: 20050408]

26. Yévenes-Briones H, Caballero FF, Struijk EA, et al. Diet Quality and the Risk of Impaired Speech Reception Threshold in Noise: The UK Biobank cohort. Ear Hear 2021;43(2):361–69. doi: 10.1097/aud.0000000000001108 [published Online First: 20210727]

27. Kant AK, Graubard BI. A comparison of three dietary pattern indexes for predicting biomarkers of diet and disease. Journal of the American College of Nutrition 2005;24(4):294–303. doi: 10.1080/07315724.2005.10719477 [published Online First: 2005/08/12]

28. Fung TT, Chiuve SE, McCullough ML, et al. Adherence to a DASH-style diet and risk of coronary heart disease and stroke in women. Arch Intern Med 2008;168(7):713–20. doi: 10.1001/archinte.168.7.713

29. Chiuve SE, Fung TT, Rimm EB, et al. Alternative dietary indices both strongly predict risk of chronic disease. The Journal of nutrition 2012;142(6):1009–18. doi: 10.3945/jn.111.157222 [published Online First: 2012/04/20]

30. Miller KL, Alfaro-Almagro F, Bangerter NK, et al. Multimodal population brain imaging in the UK Biobank prospective epidemiological study. Nat Neurosci 2016;19(11):1523–36. doi: 10.1038/nn.4393 [published Online First: 2016/10/28]

31. Alfaro-Almagro F, Jenkinson M, Bangerter NK, et al. Image processing and Quality Control for the first 10,000 brain imaging datasets from UK Biobank. NeuroImage 2018;166:400–24. doi: 10.1016/j.neuroimage.2017.10.034 [published Online First: 2017/10/29]

32. Tian Q, Pilling LC, Atkins JL, et al. The relationship of parental longevity with the aging brain-results from UK Biobank. GeroScience 2020;42(5):1377–85. doi: 10.1007/s11357-020-00227-8 [published Online First: 2020/07/17]

33. Parra-Soto S, Petermann-Rocha F, Boonpor J, et al. Combined association of general and central obesity with incidence and mortality of cancers in 22 sites. Am J Clin Nutr 2021;113(2):401–09. doi: 10.1093/ajcn/nqaa335 [published Online First: 2021/01/01]

34. Chudasama YV, Khunti K, Gillies CL, et al. Healthy lifestyle and life expectancy in people with multimorbidity in the UK Biobank: A longitudinal cohort study. PLoS medicine 2020;17(9):e1003332. doi: 10.1371/journal.pmed.1003332 [published Online First: 2020/09/23]

35. Palpatzis E, Bass N, Jones R, et al. Longitudinal association of apolipoprotein E and sleep with incident dementia. Alzheimers Dement 2021 doi: 10.1002/alz.12439 [published Online First: 2021/09/04]

36. Gauci S, Young LM, Arnoldy L, et al. Dietary patterns in middle age: effects on concurrent neurocognition and risk of age-related cognitive decline. Nutr Rev 2021 doi: 10.1093/nutrit/nuab047 [published Online First: 2021/08/16]

37. van den Brink AC, Brouwer-Brolsma EM, Berendsen AAM, et al. The Mediterranean, Dietary Approaches to Stop Hypertension (DASH), and Mediterranean-DASH Intervention for Neurodegenerative Delay (MIND) Diets Are Associated with Less Cognitive Decline and a Lower Risk of Alzheimer’s Disease-A Review. Adv Nutr 2019;10(6):1040–65. doi: 10.1093/advances/nmz054 [published Online First: 2019/06/19]

38. Akbaraly TN, Singh-Manoux A, Dugravot A, et al. Association of Midlife Diet With Subsequent Risk for Dementia. JAMA 2019;321(10):957–68. doi: 10.1001/jama.2019.1432 [published Online First: 2019/03/13]

39. Scarmeas N, Luchsinger JA, Schupf N, et al. Physical activity, diet, and risk of Alzheimer disease. JAMA 2009;302(6):627–37. doi: 10.1001/jama.2009.1144 [published Online First: 2009/08/13]

40. Scarmeas N, Stern Y, Tang MX, et al. Mediterranean diet and risk for Alzheimer’s disease. Ann Neurol 2006;59(6):912–21. doi: 10.1002/ana.20854 [published Online First: 2006/04/20]

41. Gu Y, Luchsinger JA, Stern Y, et al. Mediterranean diet, inflammatory and metabolic biomarkers, and risk of Alzheimer’s disease. J Alzheimers Dis 2010;22(2):483–92. doi: 10.3233/JAD-2010-100897 [published Online First: 2010/09/18]

42. Filippini T, Adani G, Malavolti M, et al. Dietary Habits and Risk of Early-Onset Dementia in an Italian Case-Control Study. Nutrients 2020;12(12) doi: 10.3390/nu12123682 [published Online First: 2020/12/03]

43. de Crom T, Ikram MA, Voortman T. MIND diet and risk of dementia in a populationLJbased cohort. Alzheimer’s & Dementia 2021;17(S10) doi: 10.1002/alz.055901

44. Thomas A, Féart C, Helmer C, et al. Association of a MIND diet with the risk of dementia and brain structure in a French older population. Alzheimer’s & Dementia 2021;17(S10) doi: 10.1002/alz.050386

45. Olsson E, Karlstrom B, Kilander L, et al. Dietary patterns and cognitive dysfunction in a 12-year follow-up study of 70 year old men. J Alzheimers Dis 2015;43(1):109–19. doi: 10.3233/JAD-140867 [published Online First: 2014/07/27]

46. Wagner M, Dartigues JF, Samieri C, et al. Modeling Risk-Factor Trajectories When Measurement Tools Change Sequentially During Follow-up in Cohort Studies: Application to Dietary Habits in Prodromal Dementia. Am J Epidemiol 2018;187(4):845–54. doi: 10.1093/aje/kwx293 [published Online First: 2017/10/12]

47. Jack CR, Knopman DS, Jagust WJ, et al. Tracking pathophysiological processes in Alzheimer’s disease: an updated hypothetical model of dynamic biomarkers. The Lancet Neurology 2013;12(2):207–16. doi: 10.1016/s1474-4422(12)70291-0

48. Pelletier A, Barul C, Feart C, et al. Mediterranean diet and preserved brain structural connectivity in older subjects. Alzheimers Dement 2015;11(9):1023–31. doi: 10.1016/j.jalz.2015.06.1888 [published Online First: 2015/07/21]

49. Gardener H, Scarmeas N, Gu Y, et al. Mediterranean diet and white matter hyperintensity volume in the Northern Manhattan Study. Arch Neurol 2012;69(2):251–6. doi: 10.1001/archneurol.2011.548 [published Online First: 2012/02/15]

50. Macpherson H, McNaughton SA, Lamb KE, et al. Associations of Diet Quality with Midlife Brain Volume: Findings from the UK Biobank Cohort Study. J Alzheimers Dis 2021;84(1):79–90. doi: 10.3233/JAD-210705 [published Online First: 2021/09/07]

51. Chen C, Hayden KM, Kaufman JD, et al. Adherence to a MIND-Like Dietary Pattern, Long-Term Exposure to Fine Particulate Matter Air Pollution, and MRI-Based Measures of Brain Volume: The Women’s Health Initiative Memory Study-MRI. Environ Health Perspect 2021;129(12):127008. doi: 10.1289/EHP8036 [published Online First: 2021/12/24]

52. Melo van Lent D, O’Donnell A, Beiser AS, et al. Mind Diet Adherence and Cognitive Performance in the Framingham Heart Study. J Alzheimers Dis 2021;82(2):827–39. doi: 10.3233/JAD-201238 [published Online First: 2021/06/08]

53. Akbaraly T, Sexton C, Zsoldos E, et al. Association of Long-Term Diet Quality with Hippocampal Volume: Longitudinal Cohort Study. Am J Med 2018;131(11):1372–81 e4. doi: 10.1016/j.amjmed.2018.07.001 [published Online First: 2018/07/30]

54. Titova OE, Ax E, Brooks SJ, et al. Mediterranean diet habits in older individuals: associations with cognitive functioning and brain volumes. Exp Gerontol 2013;48(12):1443–8. doi: 10.1016/j.exger.2013.10.002 [published Online First: 2013/10/16]

55. Daviet R, Aydogan G, Jagannathan K, et al. Associations between alcohol consumption and gray and white matter volumes in the UK Biobank. Nat Commun 2022;13(1):1175. doi: 10.1038/s41467-022-28735-5 [published Online First: 2022/03/06]

56. Geil CR, Hayes DM, McClain JA, et al. Alcohol and adult hippocampal neurogenesis: promiscuous drug, wanton effects. Prog Neuropsychopharmacol Biol Psychiatry 2014;54:103–13. doi: 10.1016/j.pnpbp.2014.05.003 [published Online First: 2014/05/21]

57. Kaye JA, Swihart T, Howieson D, et al. Volume loss of the hippocampus and temporal lobe in healthy elderly persons destined to develop dementia. Neurology 1997;48(5):1297–304. doi: 10.1212/wnl.48.5.1297 [published Online First: 1997/05/01]

58. Jacobs HI, Van Boxtel MP, Jolles J, et al. Parietal cortex matters in Alzheimer’s disease: an overview of structural, functional and metabolic findings. Neurosci Biobehav Rev 2012;36(1):297–309. doi: 10.1016/j.neubiorev.2011.06.009 [published Online First: 2011/07/12]

59. Wolters FJ, Chibnik LB, Waziry R, et al. Twenty-seven-year time trends in dementia incidence in Europe and the United States: The Alzheimer Cohorts Consortium. Neurology 2020;95(5):e519–e31. doi: 10.1212/WNL.0000000000010022 [published Online First: 2020/07/03]

60. Wilkinson T, Schnier C, Bush K, et al. Identifying dementia outcomes in UK Biobank: a validation study of primary care, hospital admissions and mortality data. Eur J Epidemiol 2019;34(6):557–65. doi: 10.1007/s10654-019-00499-1 [published Online First: 2019/02/27]

